# Quantifying economic returns to guide antimicrobial stewardship scale-up policy in low-resource settings

**DOI:** 10.64898/2026.08.06.26359836

**Authors:** Jung-Seok Lee, Kshitij Karki, Phonevilai Santisouk, Yunjin Yum, Wongyeong Choi, Rakchya Amatya, Bisekha Jaiswal, Geunhyeog Jang, Jaewoong Lee, Palada Souvanhnavong, Khaennakhone Salodchanar, Shrija Thapa, Naphaphone Khathtiyavong, Nilam Kumari Singh, Soulivanh Phanthavong, Loungnilanh Manivanh, Usha Tandukar, Rattanaxay Phetsouvanh, Deepak C. Bajracharya, Karishma Malla Vaidya, Khamsay Detleuxay, Sanjit Shrestha, Vangnakhone Dittaphong, Nastu Sharma, Florian Marks

## Abstract

Antimicrobial resistance is a growing threat to health systems, but stewardship programs must compete for funding with many urgent health priorities in low- and middle-income countries. Evidence that quantifies not only effectiveness but also economic value is therefore essential for policy and budget decisions. We evaluated targeted antimicrobial stewardship programs in four tertiary hospitals in Nepal and Laos using interrupted time-series analyses of antibiotic use, combined with micro-costing to estimate benefit–cost ratios. Stewardship was associated with immediate reductions in antibiotic use across the three Nepal hospitals, whereas effects in Laos were more heterogeneous. Economic returns were positive across sites, with the largest returns observed in the private hospital in Nepal. Here, we show that pragmatic, ward-focused stewardship can reduce antibiotic use and generate measurable economic value in resource-constrained hospital settings, supporting its prioritization as a scalable investment for antimicrobial resistance control.

## Background

Antimicrobial resistance (AMR) is an escalating global health threat that undermines the effectiveness of antibiotics and other antimicrobial agents and complicates the management of common infections. Recent global estimates indicate that bacterial AMR was associated with millions of deaths in 2019, with a substantial share occurring in low-resource settings where access to timely diagnostics and effective second-line therapies may be constrained [1, 2]. At the same time, antimicrobial agents remain essential to modern health systems—supporting safe surgery, cancer chemotherapy, neonatal care, and intensive care—so preserving their effectiveness is a public good with long-term benefits that extend well beyond individual patients [3].

Beyond morbidity and mortality, AMR imposes substantial economic costs through longer hospital stays, more intensive diagnostics and treatment, higher use of costly reserve agents, and productivity losses [4-6]. These costs accrue to patients, providers, and payers, increasing out-of-pocket spending and straining hospital budgets. In low- and middle-income countries (LMICs), where health systems often face limited fiscal space and multiple competing priorities—such as maternal and newborn health, tuberculosis, human immunodeficiency virus, malaria, and non-communicable diseases—decision makers must frequently prioritize interventions that deliver measurable health and financial value in the near term while also protecting long-term system resilience [3, 4].

Inappropriate and excessive antimicrobial use in health care is a key, modifiable driver of AMR, and optimizing prescribing is therefore central to national and global AMR strategies [3, 7]. Antimicrobial stewardship programs (ASPs) seek to improve antibiotic selection, dosing, route, and duration through coordinated interventions such as prescriber education, guideline implementation, audit-and-feedback, and review of ongoing therapy [8, 9]. Evidence syntheses indicate that stewardship can reduce inappropriate prescribing and may lower downstream harms such as health care–associated infections without adversely affecting clinical outcomes when appropriately implemented [10-12].

However, stewardship activities require upfront investments (e.g., staff time, training, and audit-and-feedback systems), and their benefits may be realized through multiple pathways that are not always captured in routine monitoring data. As a result, policy decisions about initiating, scaling, and sustaining ASPs are often made under uncertainty—particularly in LMIC settings where program funding must compete with other urgent health needs and where the economic case for AMR interventions is still being developed [3-5, 13]. Economic evaluations and models are therefore critical for translating changes in antibiotic use into decision-relevant metrics, so that stewardship can be compared against alternative uses of limited resources and incorporated into budgeting and financing plans [7, 14]. Evidence that demonstrates both effectiveness and value for money can also help maintain institutional and political commitment over time, which is essential for AMR control because gains can erode when stewardship activities are discontinued or deprioritized.

To address these decision needs, the current study evaluated the impact of ASP implementation across four tertiary hospitals in Nepal and Laos using a quasi-experimental study design with econometric modeling framework. These impact estimates were combined with micro-costing of ASP implementation inputs to quantify the economic return of stewardship. By jointly reporting (i) observed changes in antibiotic consumption/exposure and (ii) the economic value of those changes relative to implementation costs, the study provides pragmatic, field-based evidence that can support hospital managers and national stakeholders in making the economic case for maintaining and scaling stewardship activities amid constrained budgets and competing priorities.

## Methods

### Site selection

Four tertiary-level health facilities were selected for this study in Nepal and Laos. In Nepal, Manmohan Memorial Medical College and Teaching Hospital (Manmohan) is a 300-bed general hospital in Kathmandu. There are a total of 507 staff approximately, managing an average of 600–800 outpatient visits daily (157,307 outpatient department visits annually), 100 inpatient treatments per day (8,127 per year), and 12,749 emergency cases annually. Baidya & Banskota (B&B) Hospital in Lalitpur operates with a team of 120 doctors and over 800 staff, handling more than 90,000 outpatient registrations, over 9,000 inpatient admissions, and approximately 14,000 surgeries each year, with emergency registrations exceeding 9,000 cases annually. Paropakar Maternity and Women’s Hospital (Paropakar) in Kathmandu is a 415-bed facility with 336 beds allocated for inpatient care (241 obstetrics, 61 gynecology, and 34 newborns) and admits an average of 2,100 patients monthly. Paropakar is staffed by a multidisciplinary team of professionals, including 54 doctors, 172 nurses, 40 paramedical staff, 82 administrative/finance personnel, 247 support staff, and 27 others. In Laos, Setthathirath Hospital (STH) is centrally located in Vientiane and has 250 beds distributed across 16 wards and serves approximately 20,000 inpatients and 90,000 outpatients, making it one of the largest and most comprehensive healthcare facilities in the country. STH is recognized for advanced medical and surgical services, provides care to patients from across the region, and is evolving into a leading teaching hospital with expanded medical education and training opportunities for healthcare professionals.

### Study design and setting

A multi-site, quasi-experimental study was conducted using an interrupted time-series (ITS) design to evaluate the impact of an ASP. Hospital-level time-series outcomes were constructed for July 2024–December 2025 for the three Nepal hospitals. At STH, the study period was extended to March 2024–February 2026 to increase the number of monthly observations available for analysis and to support more stable time-series estimation. Pre-intervention and post-intervention trends were compared with the intervention time point defined at the ASP end point. Primary utilization metrics included defined daily doses (DDD) per 100 bed-days and days of therapy (DOT) per 1,000 patient-days.

### Intervention (ASP)

The ASP was implemented in all four study facilities for approximately 1.5–2 months (March 2025–April 2025). Given the short implementation period and the need to preserve interpretability for the ITS analysis, the intervention was focused on one to two selected wards per health facility. Core ASP components included prescriber education and training; review and reinforcement of antibiotic prescribing principles and locally applicable guidance; structured review of ongoing antibiotic therapy with recommendations to optimize agent selection, dose, route, and duration (including de-escalation when appropriate); and promotion of culture- and susceptibility-informed prescribing when microbiology results were available. At STH in Laos, in-person ASP training was relatively short; training was delivered on site for less than two weeks by a visiting physician, and the remaining training and support were provided virtually. For the ITS models, the intervention end point was defined as the time point at which the ASP implementation concluded; this end point was used to demarcate the pre-intervention and post-intervention periods.

### Study population and eligibility criteria

The unit of analysis for descriptive summaries was the inpatient admission (i.e., patient-level hospitalization record). Eligible records were restricted to patients admitted to the inpatient department (IPD) who were aged 15 years or older, who received antimicrobials for ≥48 hours, and who had available antimicrobial susceptibility test results.

### Microbiology and susceptibility testing

Clinical specimens submitted for routine diagnostic testing during hospitalization were included as available (e.g., blood, urine, respiratory specimens, wound/pus swabs, and other site-specific samples). Pathogens were identified by each hospital laboratory using standard routine methods, and organism identification and susceptibility results were extracted from laboratory records. Antimicrobial susceptibility testing was performed at all four facilities using the disk-diffusion method, and results were recorded using standard susceptibility categories (susceptible, intermediate, and resistant). For the purposes of analysis, isolates categorized as intermediate or resistant were classified as resistant, unless otherwise specified.

### Econometric modeling framework

The impact of the ASP was assessed using ITS regression models applied to monthly antibiotic utilization outcomes, including DDD per 100 patient-days and DOT per 1,000 patient-days. For each hospital and outcome, segmented regression models were fit to estimate (i) the baseline level at the start of the series (intercept), (ii) the pre-intervention trend (slope), (iii) the immediate level change at the ASP end point, and (iv) the post-intervention trend. The intervention was coded as a step indicator at the ASP end point and a post-intervention time term was included to capture changes in slope. Residual autocorrelation was examined using standard autocorrelation diagnostics (Supplementary Table 1). To support valid statistical inference in the presence of potential serial correlation and heteroskedasticity, Newey–West heteroskedasticity- and autocorrelation-consistent (HAC) standard errors were reported using 12 lags.

### Benefit-Cost Ratio (BCR)

The benefit–cost ratio (BCR) was estimated by combining ITS regression results with site-specific costing data. For each hospital and outcome, fitted segmented regression coefficients were used to generate observed post-intervention trajectories and counterfactual trajectories. Program benefits were quantified as the cumulative reduction in antibiotic use over the post-intervention period, calculated as the area between the observed and counterfactual curves (i.e., the difference in area under the curve) and then converted into monetary savings using each of the respective unit antibiotic cost (cost per DDD or DOT). Total ASP implementation costs were estimated per site using a micro-costing approach, with resources itemized and valued (e.g., personnel time for training and audit-and-feedback, coordination time, consumables and training materials, and travel/accommodation where applicable). The BCR was calculated as total monetary benefits divided by total ASP implementation cost. The 95% confidence intervals for BCR were estimated considering the nonlinear combinations of parameter estimates (monetary benefits to implementation costs).

### Ethical considerations

Ethical approval was obtained from the Institutional Review Board of the International Vaccine Institute (IVI-IRB) and from the relevant institutional and/or national ethics review committees in Nepal and Laos. Because the study relied on routinely collected clinical, pharmacy, and laboratory data and posed minimal risk to participants, requirements for individual informed consent were waived by the approving committees.

## Results

Descriptive statistics are shown in Table 1. A total of 2,233, 1,939, 2,686, and 688 patient-level medical records were extracted from B&B, Manmohan, Paropakar, and STH, respectively. The average patient age at each facility ranged from 53 to 55 years, except at Paropakar, where most patients were admitted for labor and delivery. Consequently, both the proportion of patients with at least one identified pathogen and the rate of antibiotic resistance were the lowest at Paropakar compared to the rest of the health facilities. As anticipated, the direct medical cost (DMC) was the highest at B&B, which is a private hospital.

**Table 1.** Descriptive statistics.

| Indicators | B&B | Manmohan | Paropakar | STH |
| --- | --- | --- | --- | --- |
| N | 2,233 | 1,939 | 2,686 | 688 |
| Mean age (years) | 53 | 55 | 27 | 55 |
| Average length of stay (days) | 8 | 6 | 5 | 6 |
| Male (%) | 54 | 45 | - | 53 |
| Patients any pathogen identified (%) | 20 | 13 | 4 | 21 |
| Patients with resistance profile among those with any pathogen identified (%) | 58 | 50 | 38 | 83 |
| DMC (US\$) | 2,248 | 365 | 95 | 223 |

Figure 1 presents the distribution of pathogens identified by health facility. *Escherichia coli* was the most frequently detected organism at all sites, accounting for 23.7% (B&B), 42.8% (Manmohan), 35.6% (Paropakar), and 52.8% (STH). *Klebsiella pneumoniae* was the next most common pathogen (23.7%, 17.9%, 16.3%, and 17.6%, respectively). *Staphylococcus aureus* was identified in B&B (9.0%), Manmohan (10.5%), and STH (5.6%) but not in Paropakar, while *Streptococcus pyogenes* was rare and observed only in B&B (0.4%) and STH (1.4%).

**Figure 1.**
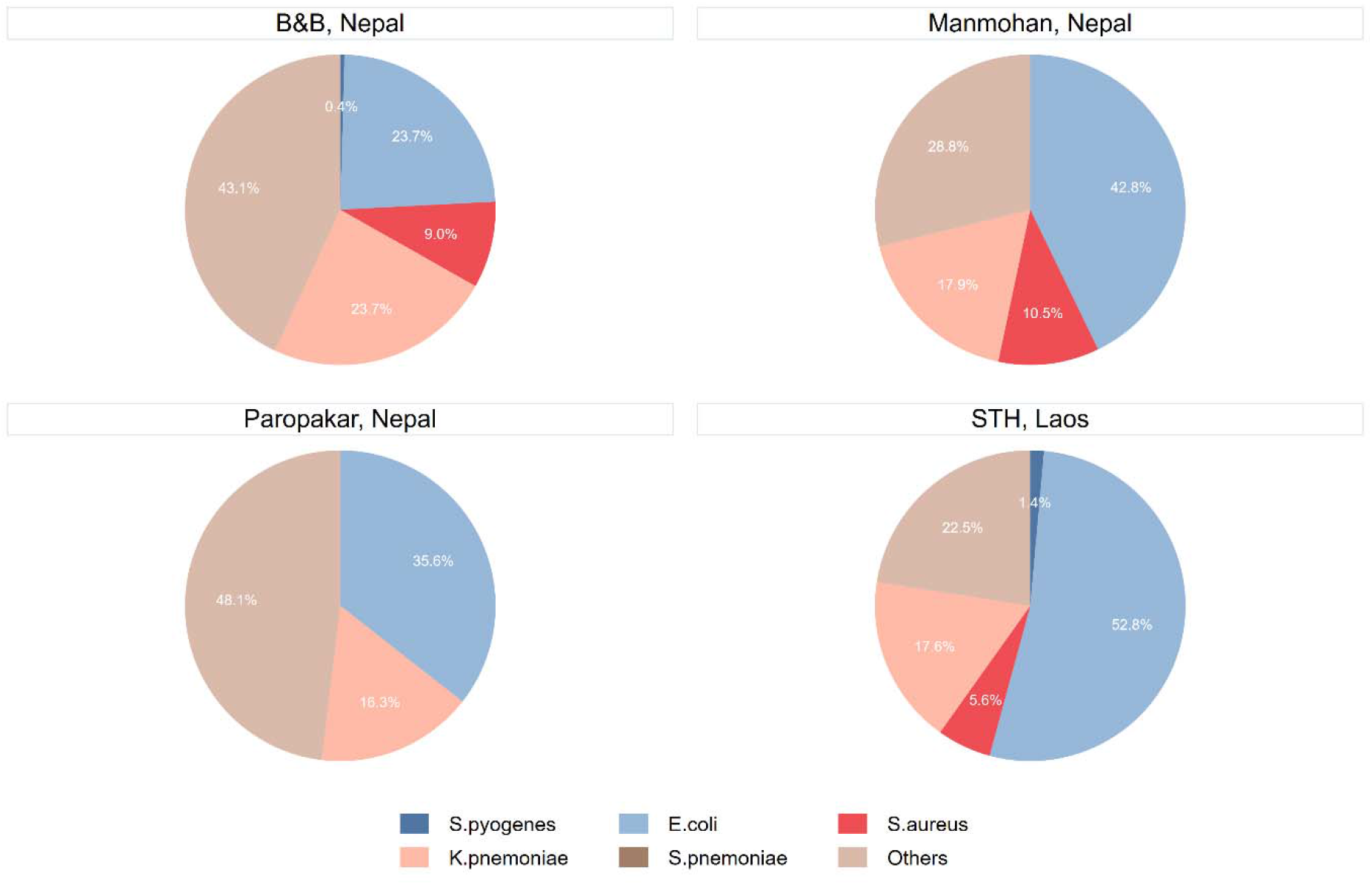
Types of pathogens identified. The antibiotic susceptibility test results are shown in Figure 2. Across four health facilities, susceptibility testing covered a total of 58 antibiotics for the patients enrolled in the current study.

**Figure 2.**
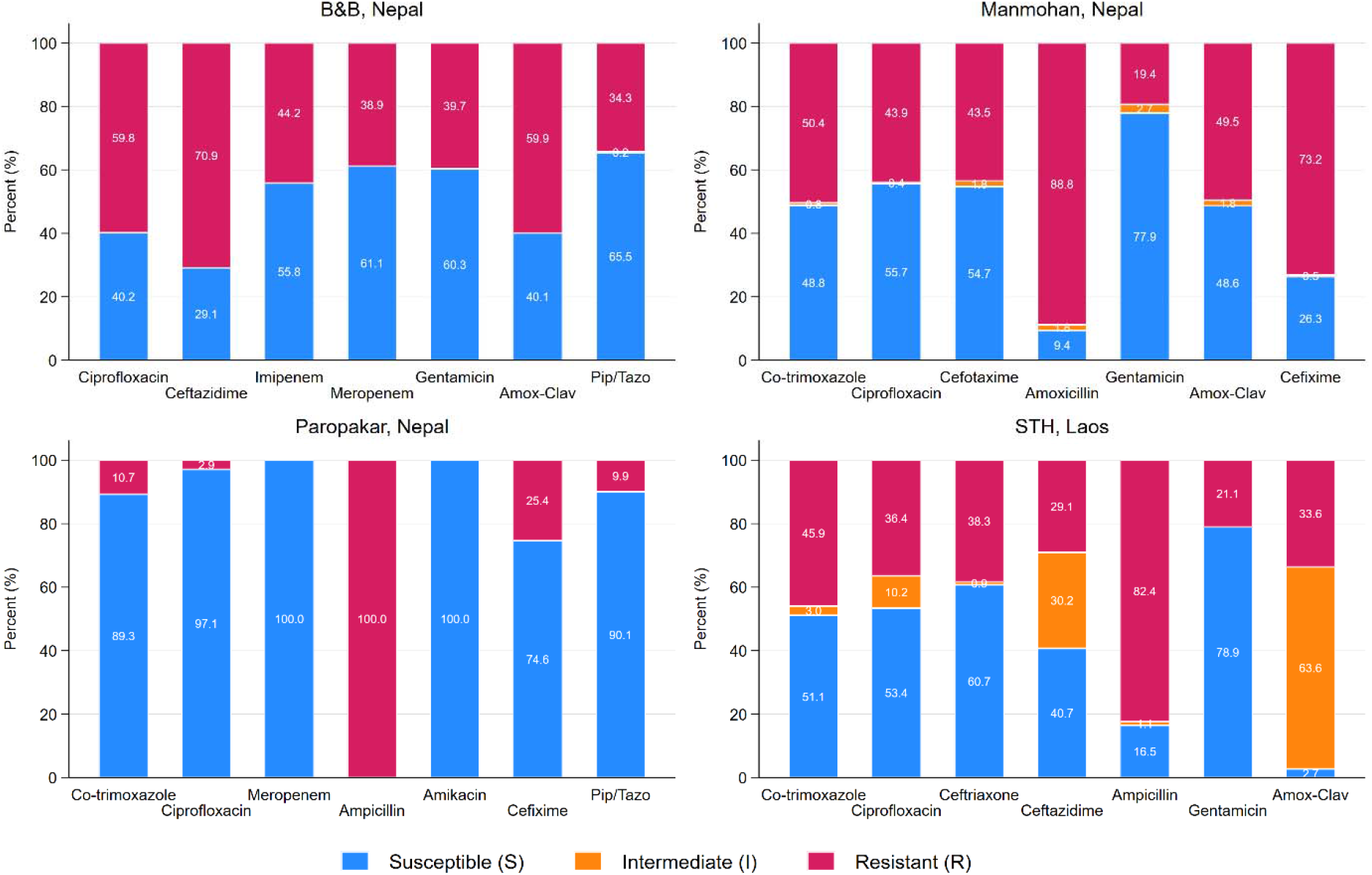
Antibiotic susceptibility test results by health facility. Out of 58 antibiotics reported during the study, the seven most frequently tested antibiotics per facility are shown in the figure.

Among the seven most commonly tested antibiotics, the level of resistance was the greatest against Ceftazidime in B&B, Amoxicillin in Manmohan, and Ampicillin in Paropakar and STH. On the other hand, several antibiotics retained relatively high susceptibility depending on the health facilities, including Piperacillin/Tazobactam in B&B, Gentamicin in Manmohan and STH, Meropenem and Amikacin in Paropakar. Notably, all pathogens tested for Meropenem and Amikacin were susceptible at Paropakar. The intermediate (I) category was most prominent in STH, especially for Amoxicillin/clavulanic acid followed by Ceftazidime. Overall, Paropakar had the lowest proportion of antibiotic-resistant pathogens among the four sites, likely reflecting its patient population, as most admissions were to the Neonatal ward for delivery rather than for infectious diseases.

Figures 3 and 4 summarize the ITS analysis of antibiotic consumption before vs after ASP implementation across the four study facilities, using two complementary utilization metrics: DDD per 100 patient-days (Figure 3) and DOT per 1,000 patient-days (Figure 4). Before the ASP end point, baseline levels differed by site, with higher DDD and DOT at B&B, intermediate levels at Manmohan and STH, and lower levels at Paropakar. Trajectories during the pre-intervention period were also heterogeneous, including increasing DDD and DOT at B&B and Paropakar and decreasing DDD at Manmohan approaching the intervention. This pattern indicates that hospitals entered the intervention period with markedly different prescribing intensities and momentum, which is important for interpreting post-intervention changes.

**Figure 3.**
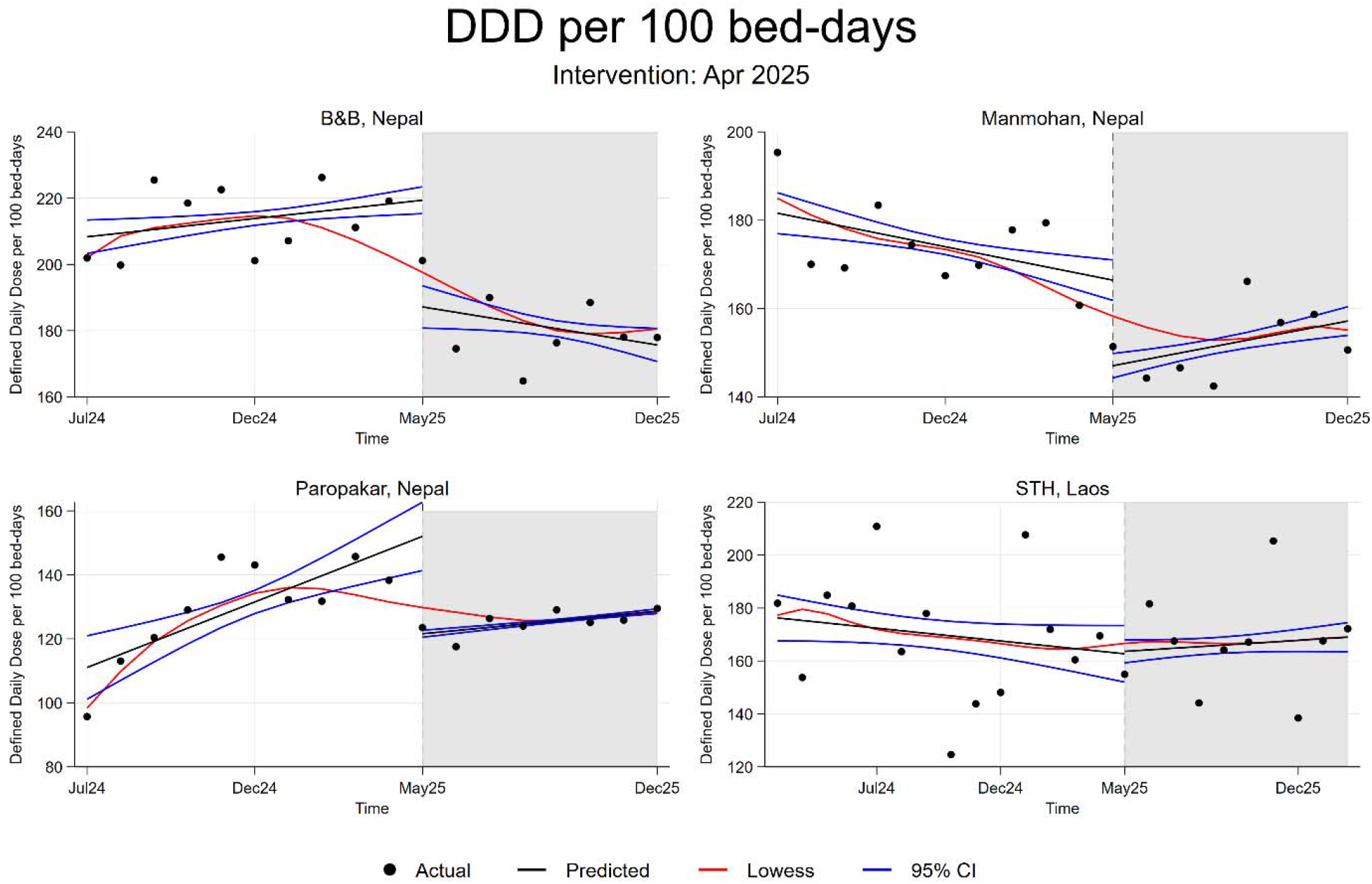
Impact of ASPs on DDD per 100 bed-days. The vertical dashed line indicates the time point when the ASP ended, thus the shaded areas show the respective indicator observed during the post-intervention period.

**Figure 4.**
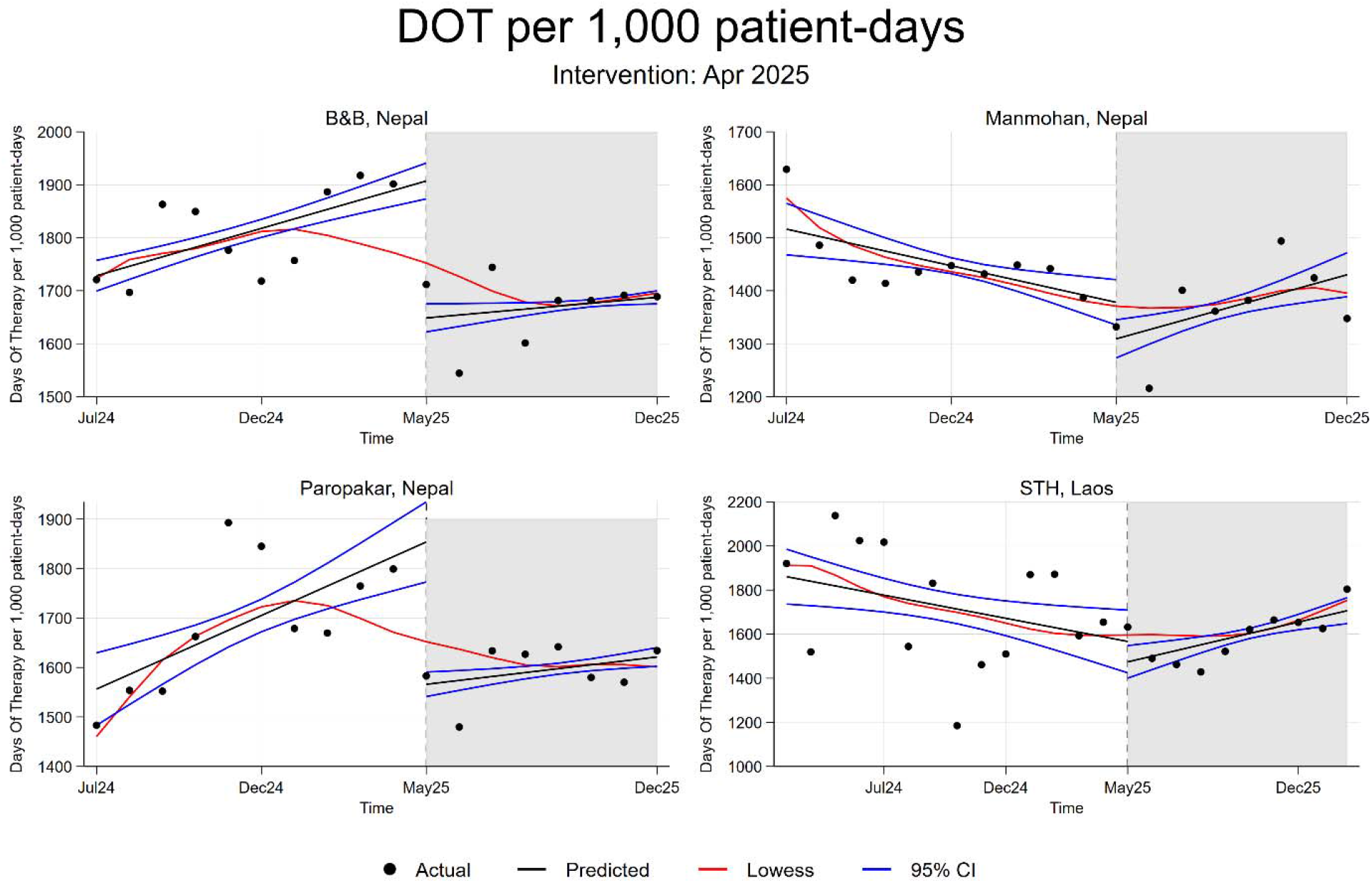
Impact of the ASPs on DOT per 1,000 patient-days. The vertical dashed line indicates the time point when the ASP ended, thus the shaded areas show the respective indicator observed during the post-intervention period.

After the ASP end point, the Nepal sites showed a visible downward shift and/or declining trend in DDD at B&B and Paropakar and an immediate reduction at Manmohan, whereas STH exhibited continued variability with a less distinct sustained change. Similarly, following the ASP end point, DOT decreased and/or stabilized at B&B and Paropakar with an immediate reduction at Manmohan, while STH remained comparatively variable and did not show an equally clear sustained reduction. Across both metrics, post-intervention patterns were more consistent with reduced or stabilized antibiotic use in the Nepal hospitals than in STH. However, it should be noted that there was a clear downward shift, followed by a modest upward shift in the Infectious Disease Adults (IDA) ward in STH (Supplementary Figure 1).

The visual patterns shown in Figures 3 and 4 are consistent with the ITS estimates in Table 2 which summarizes ITS estimates for antibiotic consumption (DDD) and exposure (DOT) by hospital, reporting baseline level, pre-intervention trend, the immediate change at the ASP end point, and the post-intervention trend. At the ASP end point, both DDD and DOT showed statistically significant immediate reductions at the three Nepal hospitals (B&B, Manmohan, and Paropakar). In contrast, STH showed no statistically significant immediate level change in either DDD or DOT at the significance level of 0.05. Post-intervention trends declined significantly at B&B and Paropakar for both metrics, whereas Manmohan and STH showed statistically significant increasing post-intervention trends, indicating site-specific heterogeneity in longer-term trajectories. Autocorrelation diagnostics (Supplementary Table 1) indicated no strong evidence of residual autocorrelation for DDD and DOT for all four health facilities (individual and joint tests generally not statistically significant) but limited potential autocorrelation at selected lags only. While these findings suggest that within-series dependence was not a prominent problem in the current study, Table 2 also reports Newey–West HAC standard errors (lag 12) to support valid inference in the presence of potential serial correlation.

**Table 2.** Regression outputs for interrupted time-series analyses.

| Indicator | Variable | B&B, Nepal |  |  | Manmohan, Nepal |  |  | Paropakar, Nepal |  |  | STH, Laos |  |  |
| --- | --- | --- | --- | --- | --- | --- | --- | --- | --- | --- | --- | --- | --- |
|  |  | Coefficients | SE | P-value | Coefficients | SE | P-value | Coefficients | SE | P-value | Coefficients | SE | P-value |
| DDD | *Cons* | 208.3 | 2.6 | 0.000 | 181.6 | 2.4 | 0.000 | 111.0 | 5.1 | 0.000 | 176.3 | 4.4 | 0.000 |
| | $\beta_1$ | 1.1 | 0.4 | 0.009 | -1.5 | 0.4 | 0.001 | 4.1 | 1.0 | 0.000 | -1.0 | 0.6 | 0.099 |
| | $\beta_2$ | -32.2 | 4.0 | 0.000 | -19.4 | 3.0 | 0.000 | -30.4 | 5.5 | 0.000 | 0.9 | 5.8 | 0.875 |
| | $\beta_3$ | -2.7 | 0.8 | 0.000 | 3.0 | 0.5 | 0.000 | -3.1 | 1.0 | 0.002 | 1.6 | 0.7 | 0.031 |
| DOT | *Cons* | 1728.3 | 14.8 | 0.000 | 1516.4 | 24.8 | 0.000 | 1556.3 | 37.4 | 0.000 | 1860.8 | 63.6 | 0.000 |
| | $\beta_1$ | 17.9 | 2.7 | 0.000 | -13.8 | 4.4 | 0.002 | 29.8 | 7.2 | 0.000 | -21.0 | 8.5 | 0.013 |
| | $\beta_2$ | -258.6 | 22.1 | 0.000 | -68.9 | 31.9 | 0.031 | -287.9 | 43.3 | 0.000 | -92.6 | 79.8 | 0.246 |
| | $\beta_3$ | -12.3 | 3.7 | 0.001 | 31.1 | 5.7 | 0.000 | -21.9 | 8.0 | 0.006 | 46.8 | 12.1 | 0.000 |

<sup>a</sup> Cons: intercept, $\beta_1$ : pre-intervention slope, $\beta_2$ : immediate level change after the intervention, and $\beta_3$ : post-intervention slope
Autocorrelation structure handled using HAC kernel (lags): Newey–West (lag 12)

Table 3 summarizes the economic returns of the ASP using the BCR. In simple terms, a BCR of 3.0 (as observed for Manmohan using DOT) means a return of $3.00 for every $1 invested in implementing the ASP. Interpreted this way, the ASP appears highly cost-beneficial at B&B, with very large returns under both DDD- and DOT-based calculations ($102 and $83, respectively). Paropakar also shows a clear positive return under both metrics, indicating that the monetary benefits associated with reduced antibiotic use outweighed implementation costs by several-fold. At Manmohan, the estimated returns are more modest but still greater than 1 although the BCR based on DDD was not statistically significant. At STH, only DOT-based results were available; these indicate a small positive return, meaning the ASP benefits were slightly greater than the implementation cost.

**Table 3.** Benefit-Cost Ratio (BCR)

| Site | DDD |  |  |  | DOT |  |  |  | ASP implementation cost |
| --- | --- | --- | --- | --- | --- | --- | --- | --- | --- |
|  | BCR | P-value | 95% CI |  | BCR | P-value | 95% CI |  |  |
|  |  |  | Lower | Upper |  |  | Lower | Upper |  |
| B&B, Nepal | 102.2 | 0.000 | 86.2 | 118.1 | 82.6 | 0.000 | 67.8 | 97.4 | \$1,204 |
| Manmohan, Nepal | 2.2 | 0.176 | -1.0 | 5.3 | 3.0 | 0.081 | -0.4 | 6.3 | \$1,229 |
| Paropakar, Nepal | 7.8 | 0.000 | 4.4 | 11.1 | 5.2 | 0.000 | 3.2 | 7.1 | \$865 |
| STH, Laos | NA | NA | NA | NA | 1.5 | 0.001 | 0.6 | 2.3 | \$1,258 |

## Discussion

In this multi-site quasi-experimental evaluation across four tertiary hospitals in Nepal and Laos, ITS analyses of DDD per 100 patient-days and DOT per 1,000 patient-days showed immediate reductions in antibiotic use in the three Nepal hospitals and more heterogeneous patterns in Laos. Differences in post-intervention slopes suggest that short, targeted stewardship can rapidly influence prescribing [10, 11], but longer-term trajectories depend on local case-mix and how well stewardship practices are sustained after initial implementation.

This study also provides an economic case for ASPs using BCRs, translating changes in antibiotic use into a decision-relevant return metric. In LMIC settings, AMR activities must compete with other urgent health priorities, and evidence on affordability and value for money can be pivotal for sustaining interventions beyond time-limited funding [4, 7, 13, 14]. Overall, BCRs indicated that the ASP was cost-beneficial in all facilities with STH showing a small positive DOT-based return in the IDA ward. The exceptionally high BCR at B&B is plausible given its private-facility context: higher medication costs increase savings from reduced use; baseline DDD (208.3) and DOT (1728.3) were comparatively high (Figures 3–4); and implementation costs were kept low by focusing on the IPD over a short period (approximately 1.5–2 months). These features also imply that returns will vary by facility depending on baseline use, unit prices, and program scope.

More broadly, the study demonstrates a feasible econometric approach for generating quantified ASP evidence in LMIC settings where rigorous evaluations remain scarce. ITS models with HAC inference leveraged routinely collected data [15]. The between-hospital heterogeneity aligns with prior evidence that ASP effects depend on baseline prescribing intensity, the feasibility of acting on microbiology results, and the intensity of audit-and-feedback [8-12]. At STH, implementation in high-acuity ICU populations likely constrained de-escalation, and ward-level patterns suggest that gains may require ongoing reinforcement. For scale-up, a phased strategy—starting in high-use wards, building local stewardship capacity, and improving microbiology turnaround and utilization monitoring—may increase the likelihood of sustained impact [7].

Several limitations should be considered. The implementation and post-intervention periods were short, limiting assessment of durability and downstream outcomes (e.g., resistance, healthcare-associated infections). In Laos, effects may have been harder to detect because one of the two wards for ASP implementation was ICU where empiric broad-spectrum therapy is common due to urgency; in-person training was brief and supplemented virtually, and the STH sample size was smaller than expected, reducing power. As with most uncontrolled ITS evaluations, unmeasured concurrent changes could have influenced trends. Finally, utilization relied on routine pharmacy data streams that differed across hospitals (dispensing/administration/prescribing), introducing potential measurement variability despite harmonized definitions.

Despite these constraints, this study adds decision-relevant evidence on both the effectiveness and economic value of hospital stewardship in LMIC settings. By combining standardized ITS impact estimation using DDD and DOT with micro-costing to estimate BCRs, the analysis strengthens the economic case for sustaining ASPs amid competing priorities and constrained budgets. The marked variation in returns—especially the high BCR in a private hospital with higher unit medication costs and higher baseline antibiotic use—underscores the importance of context-specific economic evaluation when planning stewardship scale-up.

## Data Availability

Aggregated data may be made available by the corresponding author upon reasonable request and subject to the conditions of the ethical approval.

