## Supplementary information for "Quantifying economic returns to guide antimicrobial stewardship scale-up policy in low-resource settings"

**Supplementary Table 1. Autocorrelation test outcomes**

| **Indicator** | **Lag** | **B&B, Nepal** | | | | **Manmohan, Nepal** | | | | **Paropakar, Nepal** | | | | **STH, Laos** | | | |
| --- | --- | --- | --- | --- | --- | --- | --- | --- | --- | --- | --- | --- | --- | --- | --- | --- | --- |
|  |  | **Individual lag** | | **Joint lag(s)** | | **Individual lag** | | **Joint lag(s)** | | **Individual lag** | | **Joint lag(s)** | | **Individual lag** | | **Joint lag(s)** | |
|  |  | **Chi-sq** | **P-value** | **Chi-sq** | **P-value** | **Chi-sq** | **P-value** | **Chi-sq** | **P-value** | **Chi-sq** | **P-value** | **Chi-sq** | **P-value** | **Chi-sq** | **P-value** | **Chi-sq** | **P-value** |
| DDD | 1 | 0.858 | 0.354 | 0.858 | 0.354 | 0.860 | 0.354 | 0.860 | 0.354 | 1.366 | 0.243 | 1.366 | 0.243 | 0.203 | 0.652 | 0.203 | 0.652 |
|  | 2 | 1.604 | 0.448 | 0.374 | 0.541 | 2.961 | 0.227 | 1.390 | 0.238 | 1.372 | 0.504 | 0.028 | 0.866 | 0.220 | 0.896 | 0.007 | 0.931 |
|  | 3 | 2.515 | 0.473 | 0.186 | 0.666 | 4.451 | 0.217 | 0.211 | 0.646 | 3.248 | 0.355 | 2.259 | 0.133 | 6.264 | 0.099 | 5.935 | 0.015 |
|  | 4 | 8.299 | 0.081 | 1.812 | 0.178 | 4.517 | 0.341 | 1.096 | 0.295 | 6.996 | 0.136 | 3.481 | 0.062 | 6.574 | 0.160 | 0.020 | 0.887 |
|  | 5 | 8.343 | 0.138 | 1.988 | 0.159 | 7.041 | 0.218 | 1.224 | 0.269 | 12.227 | 0.032 | 1.761 | 0.184 | 7.108 | 0.213 | 0.017 | 0.897 |
|  | 6 | 10.035 | 0.123 | 0.026 | 0.873 | 9.512 | 0.147 | 0.245 | 0.621 | 12.277 | 0.056 | 0.000 | 0.997 | 9.218 | 0.162 | 0.129 | 0.720 |
|  | 7 | 14.381 | 0.045 | 0.002 | 0.965 | 12.932 | 0.074 | 0.753 | 0.386 | 12.278 | 0.092 | 0.063 | 0.802 | 9.727 | 0.205 | 0.010 | 0.922 |
|  | 8 | 14.407 | 0.072 | 4.084 | 0.043 | 15.338 | 0.053 | 1.140 | 0.286 | 14.708 | 0.065 | 0.000 | 0.985 | 10.664 | 0.221 | 0.019 | 0.891 |
|  | 9 | 15.246 | 0.084 | 1.886 | 0.170 | 15.689 | 0.074 | 1.303 | 0.254 | 14.779 | 0.097 | 0.663 | 0.416 | 11.098 | 0.269 | 0.016 | 0.900 |
|  | 10 | 15.246 | 0.123 | 0.242 | 0.623 | 16.488 | 0.086 | 0.001 | 0.975 | 16.835 | 0.078 | 0.008 | 0.929 | 11.417 | 0.326 | 0.864 | 0.352 |
|  | 11 | 15.752 | 0.151 | 0.542 | 0.462 | 16.502 | 0.124 | 0.571 | 0.450 | 17.682 | 0.089 | 0.045 | 0.832 | 14.972 | 0.184 | 0.340 | 0.560 |
|  | 12 | 17.016 | 0.149 | 0.170 | 0.680 | 16.757 | 0.159 | 0.017 | 0.897 | 17.733 | 0.124 | 0.193 | 0.661 | 16.106 | 0.186 | 0.015 | 0.904 |
| DOT | 1 | 0.544 | 0.461 | 0.544 | 0.461 | 0.046 | 0.830 | 0.046 | 0.830 | 1.816 | 0.178 | 1.816 | 0.178 | 0.166 | 0.684 | 0.166 | 0.684 |
|  | 2 | 0.674 | 0.714 | 0.042 | 0.837 | 0.836 | 0.658 | 0.732 | 0.392 | 4.858 | 0.088 | 1.288 | 0.256 | 0.339 | 0.844 | 0.199 | 0.656 |
|  | 3 | 5.785 | 0.123 | 3.671 | 0.055 | 1.198 | 0.753 | 0.264 | 0.608 | 6.169 | 0.104 | 3.514 | 0.061 | 4.455 | 0.216 | 3.844 | 0.050 |
|  | 4 | 9.497 | 0.050 | 0.170 | 0.681 | 4.368 | 0.358 | 2.135 | 0.144 | 7.465 | 0.113 | 0.817 | 0.366 | 5.012 | 0.286 | 0.471 | 0.492 |
|  | 5 | 9.511 | 0.090 | 0.607 | 0.436 | 6.941 | 0.225 | 0.267 | 0.605 | 9.261 | 0.099 | 0.012 | 0.912 | 9.757 | 0.082 | 3.635 | 0.057 |
|  | 6 | 9.819 | 0.132 | 0.156 | 0.693 | 10.338 | 0.111 | 0.217 | 0.642 | 9.708 | 0.137 | 0.136 | 0.712 | 12.056 | 0.061 | 0.045 | 0.833 |
|  | 7 | 10.476 | 0.163 | 0.044 | 0.835 | 14.706 | 0.040 | 0.443 | 0.506 | 10.813 | 0.147 | 0.100 | 0.752 | 12.062 | 0.099 | 0.001 | 0.976 |
|  | 8 | 10.490 | 0.232 | 0.024 | 0.876 | 14.824 | 0.063 | 2.670 | 0.102 | 10.849 | 0.210 | 0.359 | 0.549 | 12.475 | 0.131 | 0.737 | 0.390 |
|  | 9 | 15.691 | 0.074 | 0.310 | 0.578 | 15.899 | 0.069 | 0.003 | 0.958 | 10.895 | 0.283 | 0.709 | 0.400 | 12.478 | 0.188 | 0.144 | 0.704 |
|  | 10 | 16.088 | 0.097 | 0.334 | 0.563 | 16.577 | 0.084 | 0.044 | 0.834 | 13.053 | 0.221 | 0.040 | 0.841 | 12.703 | 0.241 | 0.091 | 0.762 |
|  | 11 | 16.467 | 0.125 | 0.725 | 0.395 | 16.584 | 0.121 | 3.328 | 0.068 | 14.879 | 0.188 | 0.135 | 0.713 | 12.718 | 0.312 | 0.114 | 0.736 |
|  | 12 | 16.609 | 0.165 | 0.106 | 0.745 | 17.701 | 0.125 | 0.000 | 0.991 | 14.908 | 0.246 | 0.609 | 0.435 | 12.772 | 0.386 | 0.013 | 0.908 |

**Supplementary Figure 1. Impact of the ASPs on DDD and DOT by ward in STH, Laos**

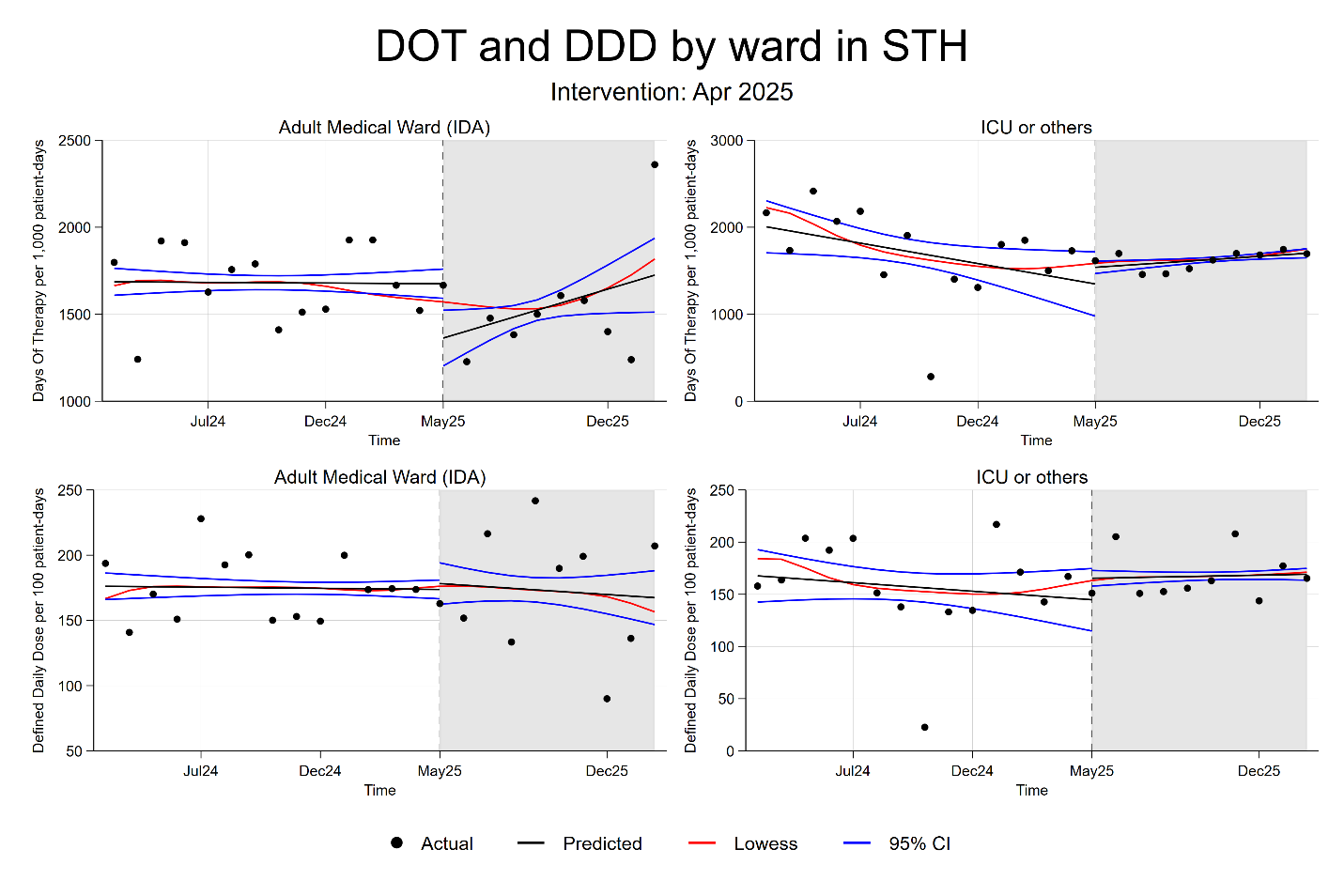
